## Supplementary Appendix for "Cost-effectiveness Analysis of Alternative Infant and Neonatal Rotavirus Vaccination Schedules in Malawi"

### Figures

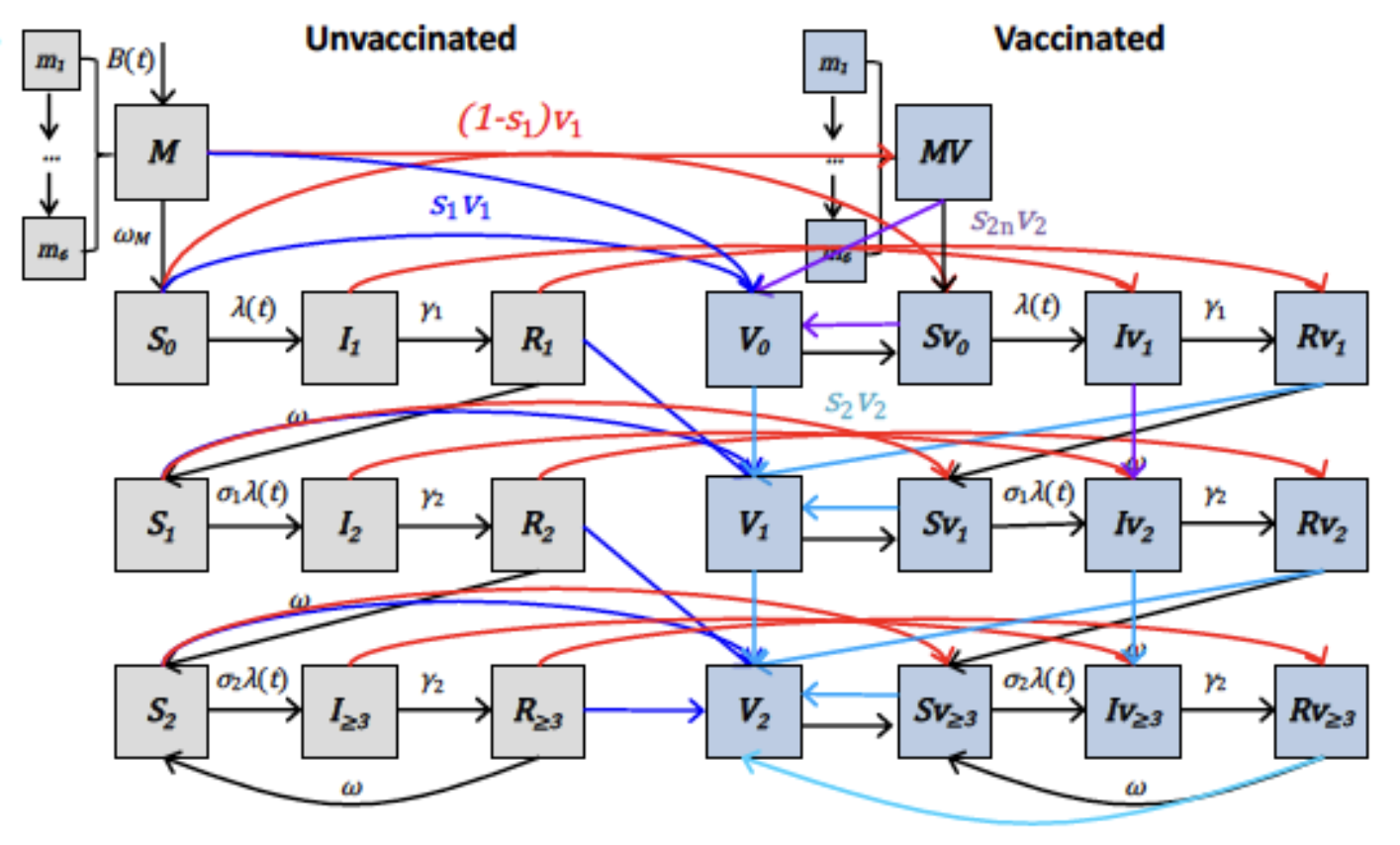

**Fig S1. Diagram of Transmission-Dynamic Model.** Compartments in grey and blue represent unvaccinated and vaccinated individuals. The arrows represent individuals transitioning between states.

**Table S1. The fixed, fitted, and estimated parameter definitions and their sources for the dynamic model.** The values in the brackets for the fitted parameters represent the 95% credible intervals. The values in the parenthesis for the estimated parameters (proportion of individuals who respond to each dose of the vaccine) represent ±20% around the mean value.

| **Parameter** | **Symbol** | **Value** | **source** |
| --- | --- | --- | --- |
| ***Fixed parameters*** |  |  |  |
| Average duration of maternal immunity | 1/*ω*_m_ | 26 weeks | (1, 2) |
| Duration of primary infection | 1/*γ1* | 1 week | (3) |
| Duration of subsequent infection | 1/*γ*_2_ | 0.5 week | (4, 5) |
| Relative risk of second infection | *σ*_1_ | 0.62 | (6, 7) |
| Relative risk of third infection | *σ*_2_ | 0.35 | (6, 7) |
| Relative infectiousness of secondary infection | *ρ*_2_ | 0.5 | (6, 7) |
| Relative infectiousness of mild/asymptomatic infections | *ρ*_≥3_ | 0.1 | (8) |
| ***Fitted parameters*** |  |  |  |
| Basic reproductive number | *R*_0_ | 78.8 (70.5-96.2) | (9) |
| Amplitude of seasonal forcing | *b* | 0.174 (0.113-0.294) | (9) |
| Seasonal offset (weeks) | *φ* | 6.9 (4.0-11.2) | (9) |
| Proportion of moderate-to-severe diarrhea cases reported | *h* | 0.017 (0.016-0.018) | (9) |
| Duration of vaccine-induced immunity (weeks) | *ω_ν_* | 45.260 (32.172-85.514) | (9) |
| ***Estimated parameters*** |  |  |  |
| **Rotarix vaccine** |  |  |  |
| Proportion who responded to the first dose | *S_C1_* | 0.527 (0.422-0.632) | (10) |
| Proportion who responded to the second dose | *S_C2_* | 0.895 (0.716-1.00) | (10) |
| Proportion who responded to the third dose | *S_C3_* | 0.895 (0.716-1.00) | (10) |
| Proportion who responded to the second dose given that they failed to respond to the first dose | *S_C2n_* | 0.117 (0.094-0.140) | (10) |
| Proportion who responded to the third dose given that they either failed to respond to first or second dose | *S_C3n_* | 0.014 (0.011-0.017) | (10) |
| **RV3-BB vaccine** |  |  |  |
| Proportion who responded to the first dose | *S_C1_* | 0.608 (0.486-0.730) | (11) |
| Proportion who responded to the second dose | *S_C2_* | 0.641 (0.513-0.769) | (11) |
| Proportion who responded to the third dose | *S_C3_* | 0.641 (0.513-0.769) | (11) |
| Proportion who responded to the second dose given that they failed to respond to the first dose | *S_C2n_* | 0.558 (0.446-0.670) | (11) |
| Proportion who responded to the third dose given that they either failed to respond to first or second dose | *S_C3n_* | 0.453 (0.362-0.544) | (11) |

**Table S2. Total number of cases, hospitalizations, DALYs, and deaths that occurred during the 2025-2035 period for each vaccine simulation.**

| Outcomes by Vaccine Schedule | | | | | | |
| --- | --- | --- | --- | --- | --- | --- |
| Total Outcomes per Strategy | | | | | | |
| Strategy | Total Cases (millions) | Non-Severe Cases (millions) | Moderate-to-Severe Cases (thousands) | Hospitalizations (thousands) | DALYs* (thousands) | Deaths |
| No vaccine | 12.4 (11.3,13.9) | 11.5 (10.4,13.0) | 905.5 (901.9,909.1) | 434.6 (418.8,451.5) | 358.1 (186.9,580.7) | 10600 (5210,17648) |
| Neonatal 1/6/10 | 9.8 (7.9,11.9) | 9.3 (7.4,11.4) | 542.2 (450.4,618.2) | 260.2 (217.8,299.9) | 223.0 (119.4,362.4) | 6400 (3125,10688) |
| Rotarix 6/10 | 10.9 (9.1,12.6) | 10.2 (8.5,11.9) | 650.9 (581.1,720.5) | 312.4 (276.8,348.8) | 263.7 (143.6,424.2) | 7600 (3688,12598) |
| Rotarix 6/10/14 | 9.9 (8.2,11.8) | 9.4 (7.7,11.2) | 548.9 (472.9,630.3) | 263.5 (226.0,303.1) | 225.4 (124.5,362.4) | 6400 (3081,10703) |
| Rotarix 6/10/40 | 9.9 (8.0,11.8) | 9.3 (7.5,11.2) | 580.4 (494.8,660.9) | 278.6 (237.0,319.3) | 236.2 (129.0,377.3) | 6800 (3258,11254) |
| All outcome means are presented with 95% Prediction Intervals | | | | | | |
| *DALYs are discounted at a rate of 3% per year | | | | | | |

**Table S3. Number of cases, hospitalizations, DALYs, and deaths averted by each strategy during the 2025-2035 period for each vaccine simulation.**

| Outcomes Averted by Vaccine Schedule | | | | | | |
| --- | --- | --- | --- | --- | --- | --- |
| Averted Outcomes per Strategy Compared to Rotarix 6/10 | | | | | | |
| Strategy | Cases Averted (millions) | Non-Severe Cases Averted (millions) | Moderate-to-Severe Cases Averted (thousands) | Hospitalizations Averted (thousands) | DALYs* Averted (thousands) | Deaths Averted |
| No vaccine | -1.5 | -1.3 | -254.6 | -122.2 | -94.4 | -3000 |
| Neonatal 1/6/10 | 1.1 | 0.9 | 108.7 | 52.2 | 40.7 | 1200 |
| Rotarix 6/10 | -- | -- | -- | -- | -- | -- |
| Rotarix 6/10/14 | 1.0 | 0.8 | 102.0 | 48.9 | 38.3 | 1200 |
| Rotarix 6/10/40 | 1.0 | 0.9 | 70.5 | 33.8 | 27.5 | 800 |
| **DALYs are discounted at a rate of 3% per year* | | | | | | |

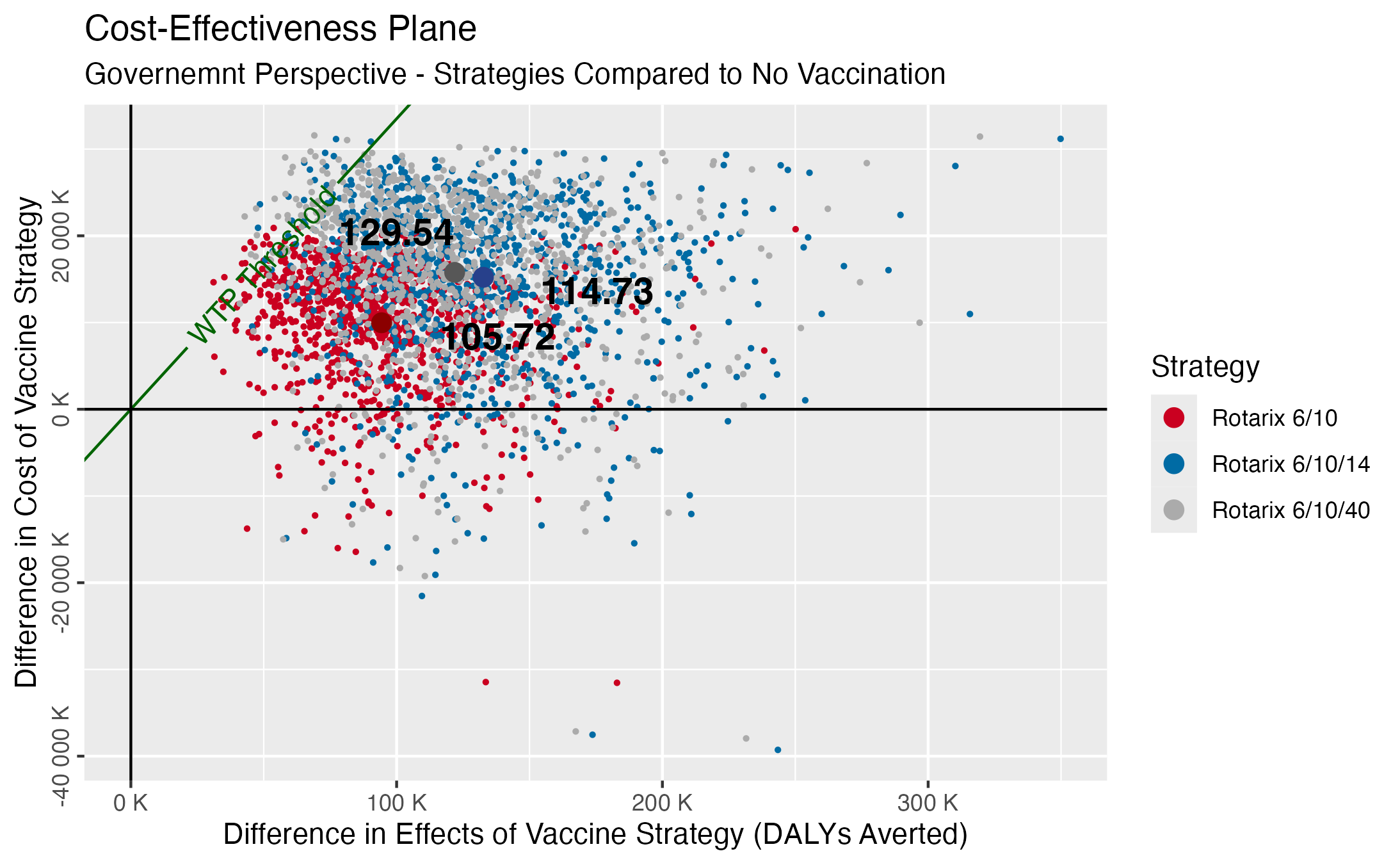

**Fig S2. Cost-effectiveness plane for the cost-effectiveness ratios of the Rotarix strategies compared to no vaccination from the government perspective.** Points represent the average cost-effectiveness ratio for each strategy. The WTP threshold is $335 per DALY averted (0.5x Malawi’s GDP per capita).

### Methods

##### Transmission Dynamic Model Description

We used an age-stratified compartmental dynamic transmission model of rotavirus (Fig S1) developed by Pitzer et al. (12). The detailed model description and fitted assumptions used to estimate the model parameters are described elsewhere in full (9, 13). To summarize, infants are born into the maternal compartment at a rate determined by Malawi's crude birth rate. In the maternal compartment, infants are protected by maternally acquired antibodies, which wane before infants become susceptible to their first rotavirus infection. Once this immunity wanes, individuals are infected at a rate determined by the force of infection, with a proportion of infected infants developing moderate-to-severe rotavirus-associated gastroenteritis (RVGE). Following infection, infants recover into a temporary immune compartment where they are protected against reinfections. After the waning of this naturally acquired temporary immunity, individuals become susceptible to their second infection at a reduced rate. Compared to primary infection, a smaller fraction develops moderate-to-severe RVGE and individuals recover at a faster rate to the temporary immune compartment. Subsequent infections follow similar transitions, but with infections mostly resulting in either mild or asymptomatic rotavirus. The population is stratified into 42 age groups (monthly intervals for infants less than two years, yearly intervals from 2-4 years, 5-year intervals from 5 to 70 years, and those above 70 years), and we assume a homogeneous mixing pattern.

To model vaccination, we assume that infants who respond after receiving a single dose of the vaccine are temporarily protected against rotavirus infection. Following the waning of vaccine-induced immunity, infants become susceptible again and can be infected with rotavirus, potentially leading to moderate-to-severe disease. However, infants who have responded to at least two doses of the vaccine are assumed to be permanently protected from moderate-to-severe RVGE and can only be infected with non-severe rotavirus following the waning of vaccine-induced immunity. The age of vaccination in the model is determined by the dosing schedule and rounded to the nearest month of age to account for realistic delays in vaccine delivery (Table S4).

**Table S4. The dosing schedules, the number of doses, and the age of vaccination.**

| **Schedule (weeks)** | **Number of doses** | **Age at vaccination (months)** |
| --- | --- | --- |
| 6/10 | 2 | 2, 3 |
| 6/10/14 | 3 | 2, 3, 4 |
| 6/10/40 | 3 | 2, 3, 9 |
| 1/6/10 | 3 | 0, 2, 3 |

##### Cost-Effectiveness Model

The analyses were run twice, once to determine the optimal strategy from the government perspective and another from the societal perspective. The government perspective included all costs paid by the government of Malawi related to medical treatment, buying vaccine doses with Gavi support, and delivering the vaccine. The societal perspective included costs to the government and direct household costs paid by individuals seeking treatment for RVGE, including treatment, transportation, and loss of productivity costs.

Here, we further describe the sources of our parameters. Some probability parameter estimates come from a series of related studies conducted in Kenya about the impact of RVGE of varying severities on the population and their health-seeking behaviors, as data from Malawi was unavailable(14, 15). Case fatality rate (CFR) estimates were derived from a recent systematic review that found hospital-based studies reported lower CFRs than community-based studies(16). The life expectancy at birth was for children <5 years old at the start of the simulation, which, along with gross domestic product (GDP) per capita, were from World Bank reports(17, 18). The cost of the Rotarix vaccine is the expected price for Gavi-eligible countries in 2025. A 2023 study in Ghana provided the estimated cost of switching from Rotarix to a different rotavirus vaccine in this analysis, which is a conservative estimate considering Ghana’s much larger health system(19-21). The inpatient and outpatient treatment costs for moderate-to-severe and non-severe RVGE were estimates from patient-level costing data in Malawi(22).

### Results

##### Societal Perspective

From the societal perspective, we predicted the current Rotarix vaccine strategy, in effect, costs about $25 million to implement and results in $83 million in treatment costs over ten years, about $7 million more in total than the cost of no vaccine strategy (Table 2). The 6/10/14 and 6/10/40 Rotarix schedules cost about $112 and $113 million respectively. The neonatal strategy was estimated to incur the least cost increase compared to all other vaccine strategies, saving about $5 million compared to the current two-dose Rotarix schedule and costing $2.5 million more than no vaccination. Switching from the 6/10 to the neonatal RV3-BB strategy saved society an estimated $1.1 million in direct household costs over ten years.

**Table S5. DALYs averted and incremental cost-effectiveness ratios for all vaccine strategies compared to the current Rotarix 6/10 schedule from the societal perspective.**

| Cost-Effectiveness Comparison of All Vaccine Strategies | | | | | |
| --- | --- | --- | --- | --- | --- |
| Societal perspective | | | | | |
| Strategy | Cost (millions) | DALYs (thousands) | Incremental Cost (millions) | DALYs Averted (thousands) | ICER ($/DALY averted) vs next best alternative |
| No vaccine | $101.1 | 358.1 | --- | --- | --- |
| Neonatal 1/6/10 | $103.7 | 223.0 | $2.6 | 135.1 | $19.25 |
| Rotarix 6/10 | $108.5 | 263.7 | $4.8 | -40.70 | **Dominated** |
| Rotarix 6/10/14 | $112.6 | 225.4 | $8.9 | -2.40 | **Dominated** |
| Rotarix 6/10/40 | $113.4 | 236.2 | $9.7 | -13.20 | **Dominated** |
| Costs reflect 2025 USD | | | | | |
| In conformity with accepted practice, all incremental costs and DALYs averted are computed compared to the next smallest, non-dominated strategy | | | | | |

Table S5 shows that from a societal perspective, all Rotarix schedules were strongly dominated by the neonatal RV3-BB vaccine. The model estimated the neonatal RV3-BB vaccine averted the most DALYs and had an even lower ICER compared to the Rotarix 6/10 strategy from the societal perspective (Fig S3). The RV3-BB vaccine strongly dominated all other strategies and remained cost-effective at all WTP thresholds. Unlike the government perspective analysis, the acceptability and frontier curves show the neonatal RV3-BB was preferred even at a WTP threshold of $19.25 per DALY (Table S5, Fig S4). We conclude that all other strategies are still expected to be strongly dominated by the RV3-BB vaccine, and the RV3-BB strategy is even more cost-effective from a societal perspective.

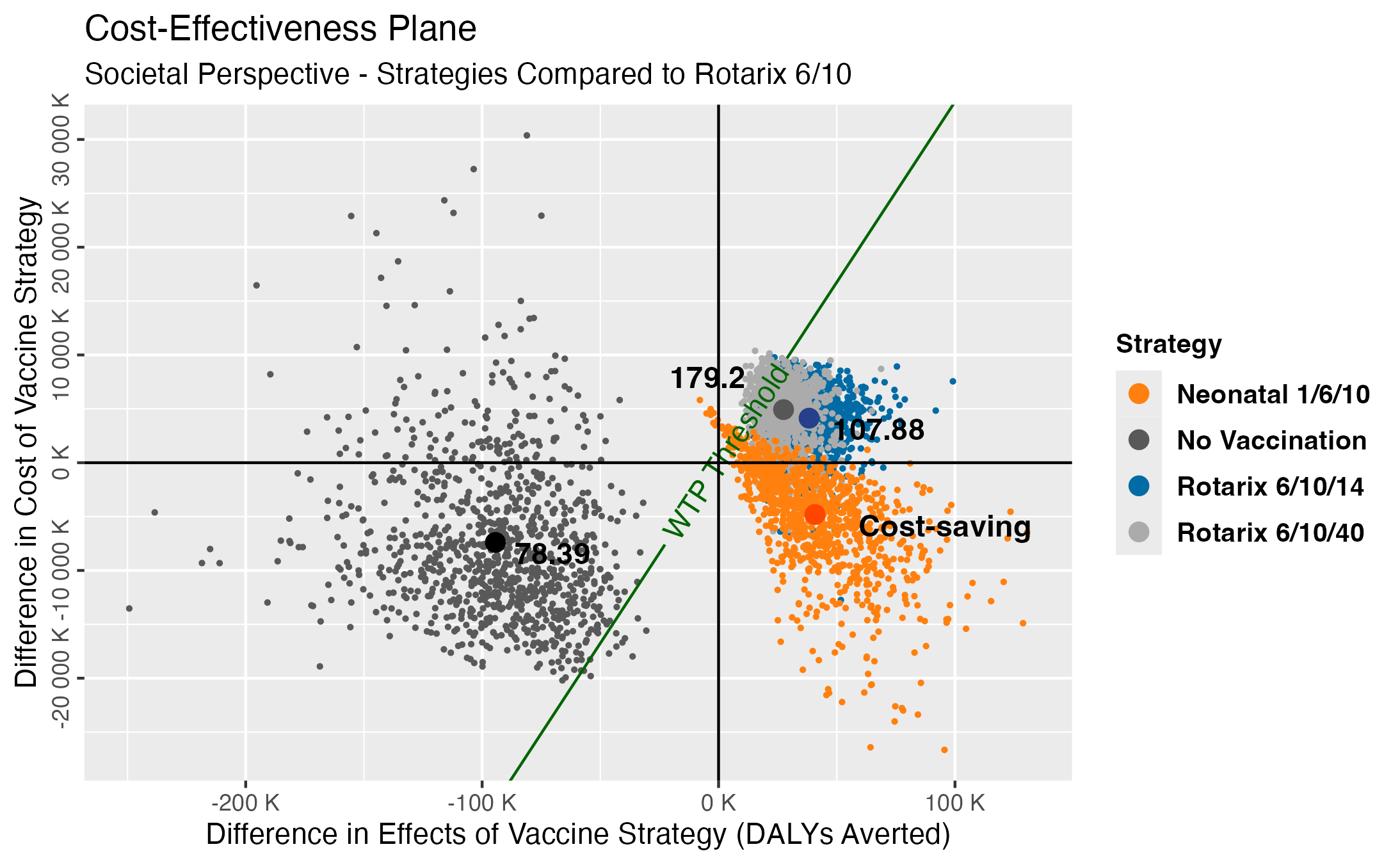

**Fig S3. Cost-effectiveness plane for the cost-effectiveness ratios of the four strategies compared to the current Rotarix 6/10 schedule from the societal perspective.** Points represent the average cost-effectiveness ratio for each strategy. The WTP threshold is $335 per DALY averted (0.5x Malawi’s GDP per capita).

The cost-effectiveness ratios compared to the Rotarix 6/10 strategy from the societal perspective followed a similar trend on the cost-effectiveness plane as those from the government perspective, except they all fell lower on the y-axis, meaning they saved more money (Fig S3). The three-dose and no-vaccine strategies were more likely acceptable at lower WTP thresholds, but the neonatal vaccine remained optimal.

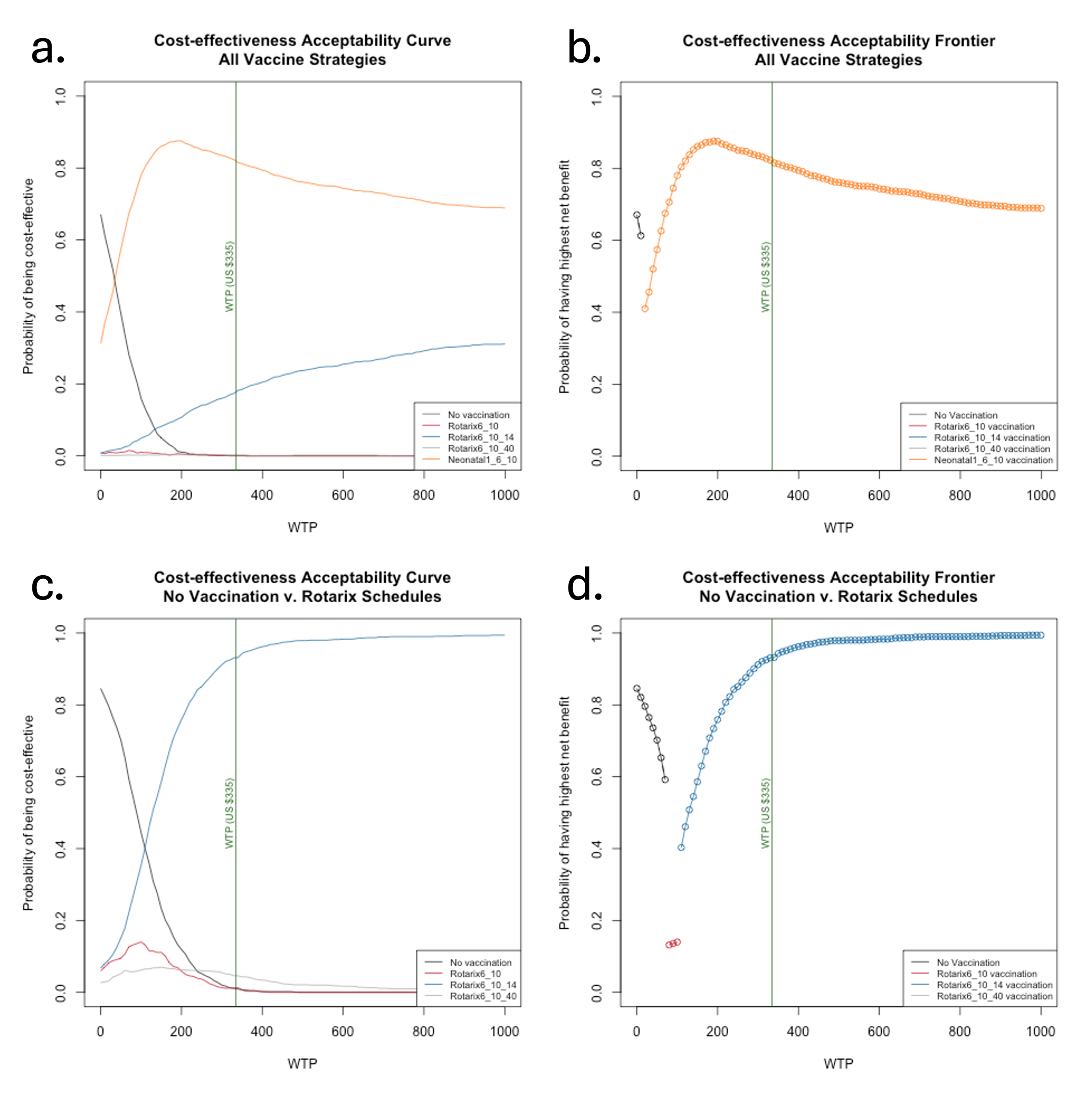

**Fig S4. Cost-effectiveness acceptability (a) curves and (b) frontier for all strategies and (c) curves and (d) frontier for currently available strategies from the societal perspective.** The vertical green line represents 0.5x Malawi’s GDP per capita.

Notable differences between the government and societal perspective analysis results appear in the cost-effectiveness ratios and the WTP thresholds at which different strategies become optimal. We predicted the average ICER for the neonatal vaccine strategy was much lower from the societal perspective, saving households an additional $1.1 million in treatment costs. The secondary analysis was also performed from a societal standpoint to determine the best Rotarix strategy without the RV3-BB vaccine. The Rotarix 6/10 strategy is cost-effective at a WTP of about $78-107, and Rotarix 6/10/14 is cost-effective at a WTP of $107 and greater (Table S6). Because the Rotarix 6/10/14 ICER is lower, from the societal perspective, this strategy was acceptable at a lower WTP (Fig S4c,d, Table S6, Fig S5). When considering all vaccines, RV3-BB was the optimal strategy overall. The sensitivity analysis determined the maximum acceptable price for one dose of RV3-BB should remain below $2.5 compared to the Rotarix 6/10 strategy and $1.90 compared to the Rotarix 6/10/14 strategy (Fig S5).

**Table S6. DALYs averted and incremental cost-effectiveness ratios for available vaccine strategies compared to no vaccination from the societal perspective.**

| Cost-Effectiveness Comparison of Available Vaccine Strategies | | | | | |
| --- | --- | --- | --- | --- | --- |
| Societal perspective | | | | | |
| Strategy | Cost (millions) | DALYs (thousands) | Incremental Cost (millions) | DALYs Averted (thousands) | ICER ($/DALY averted) vs next best alternative |
| No vaccine | $101.1 | 358.1 | --- | --- | --- |
| Rotarix 6/10 | $108.5 | 263.7 | $7.4 | 94.4 | $78.39 |
| Rotarix 6/10/14 | $112.6 | 225.4 | $4.1 | 38.3 | $107.05 |
| Rotarix 6/10/40 | $113.4 | 236.2 | $0.8 | -10.8 | **Dominated** |
| Costs reflect 2025 USD | | | | | |
| In conformity with accepted practice, all incremental costs and DALYs averted are computed compared to the next smallest, non-dominated strategy | | | | | |

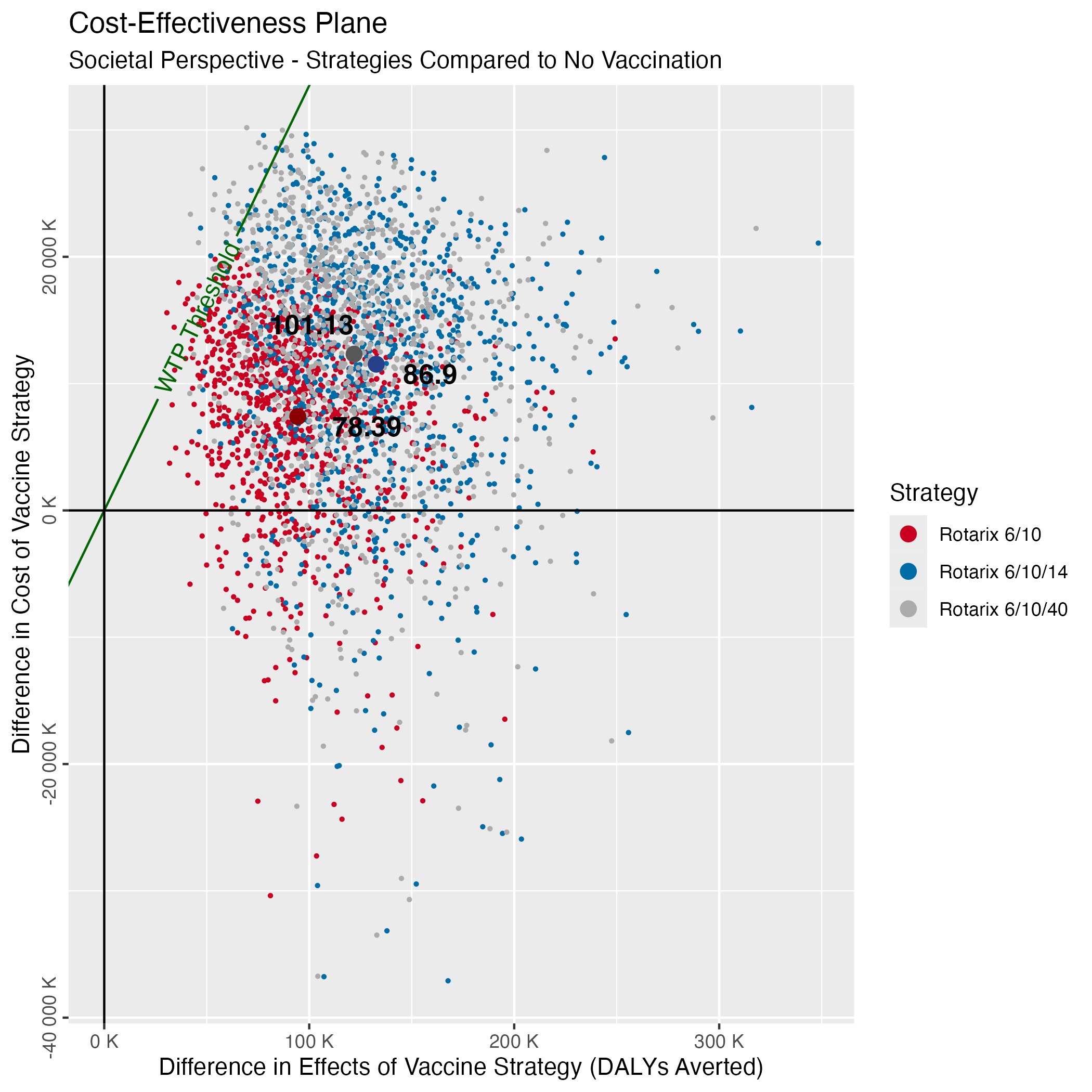

**Fig S5. Cost-effectiveness plane for the cost-effectiveness ratios of the Rotarix strategies compared to no vaccination from the societal perspective.** Points represent the average cost-effectiveness ratio for each strategy. The WTP threshold is $335 per DALY averted (0.5x Malawi’s GDP per capita).

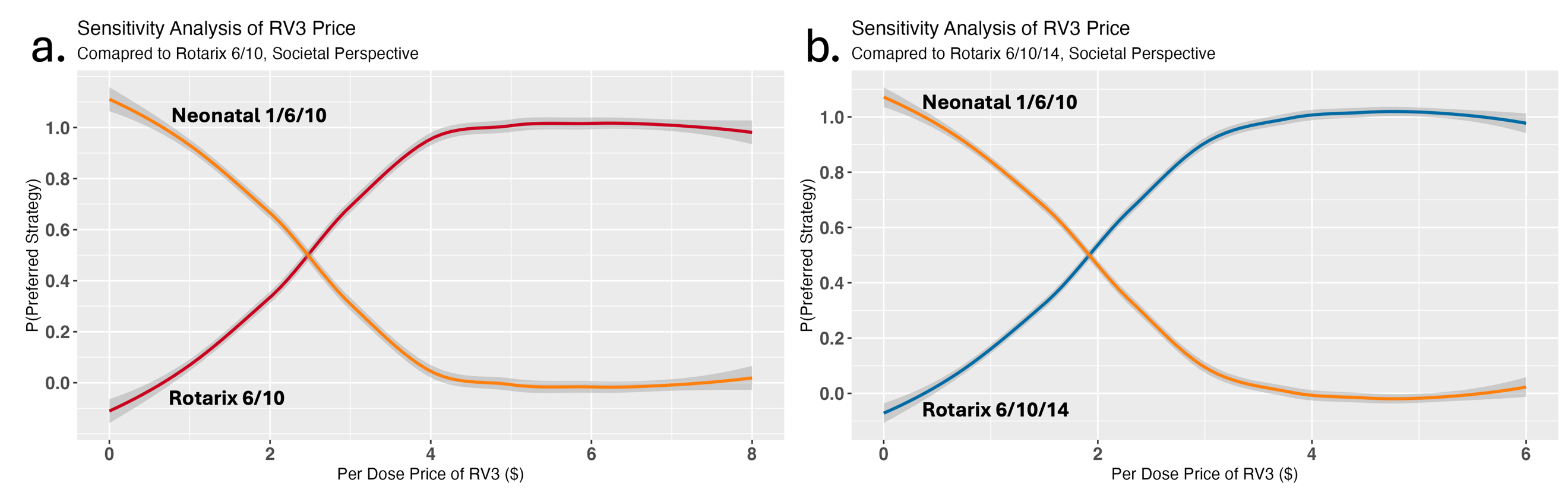

**Fig S5. Sensitivity analyses of the price per dose of the RV3-BB neonatal vaccine compared to the current Rotarix schedule and the optimal three-dose Rotarix schedule from the societal perspective.**

### Discussion

From the societal perspective, most rotavirus vaccine strategies saved about $1 million each at the household level, but only RV3-BB was estimated to save money overall compared to the current two-dose vaccine schedule. A cost-effective analysis that re-evaluated the rotavirus program in Malawi in 2018 found that the current 6/10 Rotarix schedule would avert $8 million from the healthcare perspective and $9.2 million (13% more) from the societal perspective(23). In this study, over $15 million in health costs was averted from the government perspective and about $18 million (20% more) from the societal perspective compared to no vaccination. The difference in perspectives has grown, suggesting that although costs may have increased over time, and this analysis considered additional costs, the marginal benefit to society has also increased. While there are no other apparent cost-effectiveness studies of the RV3-BB vaccine in Malawi from the societal perspective, the societal benefits of rotavirus vaccines in general, coupled with the significant life and cost-saving attributes of the neonatal vaccine, make a strong case for switching to this new vaccine if and when it becomes available.

### References

1.	Pitzer VE, Viboud Cc, Lopman BA, Patel MM, Parashar UD, Grenfell BT. Influence of birth rates and transmission rates on the global seasonality of rotavirus incidence. Journal of the Royal Society Interface. 2011;8:10.

2.	Patel M, Shane AL, Parashar UD, Jiang B, Gentsch JR, Glass RI. Oral rotavirus vaccines: how well will they work where they are needed most? J Infect Dis. 2009;200 Suppl 1(0 1):S39-48.

3.	Kambhampati A, Payne DC, Costantini V, Lopman BA. Host Genetic Susceptibility to Enteric Viruses: A Systematic Review and Metaanalysis. Clinical Infectious Diseases. 2016;62(1):8.

4.	Ultsch B, Damm O, Beutels P, Bilcke J, Bru¨ggenju¨rgen B, Gerber-Grote A, et al. Methods for Health Economic Evaluation of Vaccines and Immunization Decision Frameworks: A Consensus Framework from a European Vaccine Economics Community. PharmacoEconomics. 2015;34:17.

5.	Gelman A, Rubin DB. Inference from Iterative Simulation Using Multiple Sequences. Statistical Science. 1992;7(4):16.

6.	Velázquez FR, Matson DO, Calva JJ, Guerrero L, Morrow AL, Carter-Campbell S, et al. Rotavirus infection in infants as protection against subsequent infections. New England Journal of Medicine. 1996;335(14):7.

7.	Gladstone BP, Ramani S, Mukhopadhya I, Muliyi J, Sarkar R, Rehman AM, et al. Protective effect of natural rotavirus infection in an Indian birth cohort. New England Journal of Medicine. 2011;365(4):10.

8.	Van Effelterre T, Soriano-Gabarró M, Debrus S, Newbern EC, Gray J. A mathematical model of the indirect effects of rotavirus vaccination. Epidemiology & Infection. 2010;138(6):14.

9.	Pitzer VE, Bennett A, Bar-Zeev N, Jere KC, Lopman BA, Lewnard JA, et al. Evaluating strategies to improve rotavirus vaccine impact during the second year of life in Malawi. Science Translational Medicine. 2019;11:12.

10.	Cunliffe NA, Witte D, Ngwira BM, Todd S, Bostock NJ, Turner AM, et al. Efficacy of human rotavirus vaccine against severe gastroenteritis in Malawian children in the first two years of life: a randomised, double-blind, placebo controlled trial. Vaccine. 2012;30(1):16.

11.	Witte D, Handley A, Jere KC, Bogandovic-Sakran N, Mpakiza A, Turner A, et al. Neonatal rotavirus vaccine (RV3-BB) immunogenicity and safety in a neonatal and infant administration schedule in Malawi: a randomised, double-blind, four-arm parallel group dose-ranging study. Lancet Infectious Disease. 2022;22:11.

12.	Pitzer VE, Viboud C, Simonsen L, Steiner C, Panozzo CA, Alonso WJ, et al. Demographic variability, vaccination, and the spatiotemporal dynamics of rotavirus epidemics. Science. 2009;325(5938):5.

13.	Pitzer VE, Ndeketa L, Asare EO, Hungerford D, Lopman BA, Jere KC, et al. Impact of rotavirus vaccination in Malawi from 2012 to 2022 compared to model predictions. npj Vaccines. 2024;9(227).

14.	Omore R, O’Reilly CE, Williamson J, Moke F, Were V, Farag TH, et al. Health Care-Seeking Behavior During Childhood Diarrheal Illness: Results of Health Care Utilization and Attitudes Surveys of Caretakers in Western Kenya, 2007–2010. American Journal of Tropical Medicine. 2013;89:12.

15.	Omore R, Khagayi S, Ogwel B, Onkoba R, Ochieng JB, Juma J, et al. Rates of hospitalization and death for all-cause and rotavirus acute gastroenteritis before rotavirus vaccine introduction in Kenya, 2010–2013. BMC Infectious Diseases. 2019;19(47):11.

16.	Asare EO, Hergott D, Seiler J, Morgan B, Archer H, Wiyeh AB, et al. Case fatality risk of diarrhoeal pathogens: a systematic review and meta-analysis. International Journal of Epidemiology. 2022;00:12.

17.	Life expectancy at birth, total (years) - Malawi: World Bank; 2021 [Available from: <https://data.worldbank.org/indicator/SP.DYN.LE00.IN?locations=MW>.

18.	GDP per capita - Malawi: World Bank; 2022 [Available from: <https://data.worldbank.org/indicator/NY.GDP.PCAP.CD?locations=MW>.

19.	Owusu R, MvunduraI M, NonvignonI J, Armah G, Bawa J, Antwi-Agyei KO, et al. Rotavirus vaccine product switch in Ghana: An assessment of service delivery costs, switching costs, and cost-effectiveness. PLOS Global Public Health. 2023;3(8):17.

20.	Muula A, Dambula I, Katengeza H, Nakoma P. National and subnational coverage and other service statistics for reproductive, maternal, newborn and child health from health facility data and surveys - Malawi. Malawi Ministry of Health; 2022 13 June 2022.

21.	Niohuru I. Health Resources. SpringerBriefs in Economics: Springer; 2023. p. 87-104.

22.	Bar-Zeev N, Tate JE, Pecenka C, Chikafa J, Mvula H, Wachepa R, et al. Cost-Effectiveness of Monovalent Rotavirus Vaccination of Infants in Malawi: A Postintroduction Analysis Using Individual Patient–Level Costing Data. Clinical Infectious Diseases. 2016;62:9.

23.	Pecenka C, Debellut F, Bar-Zeev N, Anwari P, Nonvignon J, Shamsuzzaman M, et al. Re-evaluating the cost and cost-effectiveness of rotavirus vaccination in Bangladesh, Ghana, and Malawi: A comparison of three rotavirus vaccines. Vaccine. 2018;36:7.
